# The psychological impact of childbirth: Unscheduled cesarean delivery increases risk for acute stress response

**DOI:** 10.64898/2025.12.05.25340759

**Authors:** Hadas Allouche-Kam, Isha Hemant Arora, Christina T Pham, Eunice Chon, Mary Lee, Onyekachi Agwu, JiaJia Zhang, Evelyn Milavsky, Andrea G. Edlow, Francine Hughes, Scott P. Orr, Anjali J Kaimal, Sharon Dekel

## Abstract

**Introduction:** Cesarean delivery is one of the most common surgical procedures in the United States. Despite their frequency and medical necessity, the acute psychological impact of unscheduled cesarean delivery is unknown. This study aimed to assess the rates and nature of peritraumatic stress response during and shortly after unscheduled cesarean delivery, and whether acute stress predicts later mental health symptoms.

**Methods:** 1,146 patients receiving routine perinatal care at a tertiary, urban hospital were assessed for peritraumatic stress reactions during the delivery hospitalization (mean: 31 hours postpartum). A subgroup (n = 795; 69.4%) completed an assessment at approximately 1.9 months postpartum. Delivery mode, obstetric complications, prior trauma exposure, and antepartum and postpartum psychiatric symptoms were obtained from questionnaires and medical records. Chi-square tests adjusted relative risk estimates, mixed-effects models and regression analyses were used to evaluate acute stress by delivery mode, its persistence over time, and associations with postpartum mental health outcomes.

**Results:** 10.4% of participants met criteria for clinical acute stress (PDI ≥ 15). Among women undergoing unscheduled cesarean, 26.6% reported clinical stress, with higher rates observed for cesarean performed during labor (29.3%) and those with greater obstetric morbidity (ρ=0.20, p< 0.005). Compared with vaginal delivery, unscheduled cesarean delivery was associated with fourfold increased risk of acute stress (26.6% vs. 6.3%, relative risk=4.20, 95% CI: 2.92–6.05). Adjusting for demographics, primiparity, obstetric complications, labor induction, prior trauma, and antepartum mental health, unscheduled cesarean remained associated with increased risk. Stress levels among patients undergoing unscheduled cesareans was persistently elevated over time (estimate=0.39, p=0.44), while vaginal delivery was associated with a significant symptom reduction (estimate=0.52, p=0.03). Acute stress strongly predicted subsequent posttraumatic (β=0.48, p<0.001) and depressive (β=0.30, p<0.001) symptoms and maternal-infant bonding (β=0.32, p<0.001) difficulties.

**Conclusions:** A substantial proportion of women undergoing unscheduled cesarean delivery experience significant psychological stress during childbirth with enduring morbidities. Screening for acute traumatic stress during postpartum stay is warranted to optimize mental health outcomes.

**Key Points:** Childbirth can be a traumatic experience yet how it associates with delivery mode and obstetric complications is poorly understood.

In immediate postpartum assessments, 27% of women undergoing unscheduled cesarean delivery reported clinically significant acute stress using the peritraumatic distress inventory (PDI), with elevated symptoms persisting after two months and associated with maternal mental health morbidities. Compared to vaginal delivery, unscheduled cesarean was associated with a threefold increased risk, after adjusting for parity, mental health history, trauma exposure, obstetric complications, and labor induction.

Unscheduled cesarean delivery is associated with substantial acute psychological stress with enduring implications. This warrants immediate screening for birth trauma to prevent postpartum psychological morbidity.

## Introduction

Childbirth is an intense event, entailing rapid changes across multiple systems,^1–3^ leading to significant hormonal and physiological fluctuations,^4–7^ and often resulting in significant pain.^8–9^ A substantial portion of births (∼32%) in the United States are by cesarean delivery,^10^ a major surgical procedure^11–12^ that in many cases involves an unscheduled, and at times emergent, intervention.^13^ An estimated 1 in 5 women in the first delivery undergo an unscheduled cesarean delivery (CD)^14–15^ that is typically performed during active labor due to failure to progress or non-reassuring fetal assessment^14–16^ and intended to reduce maternal and neonatal morbidity and mortality.^17^

Existing data show that unscheduled CD and/or the underlying indications for the procedure can be associated with short and long-term medical implications for maternal recovery, infant health, and future pregnancies.^18–22^ However, far less is known about the psychological consequences of unscheduled CD, particularly its acute emotional impact.

Unscheduled CDs and the circumstances surrounding them may fulfill DSM-5 Criterion A for exposure to a traumatic event, involving real or perceived threats to life or serious injury, either to the self or others.^23^ The unanticipated need for major surgery especially in the context of prior physiological and emotional strain during active labor may provoke intense emotional responses and perceived traumatization.^24^ A large body of research has documented peri- traumatic stress reactions that occur during or immediately after a traumatic event across various trauma-exposed samples.^25–33^ These responses are shown to be strong predictors of subsequent posttraumatic stress disorder (PTSD),^24–34^ even more so than pre-trauma vulnerability factors.^32,35^ Understanding the magnitude of peri-traumatic stress associated with unscheduled CD may have important implications for targeted screening and implementations of clinical approaches in labor and delivery and postpartum units to enhance maternal psychological resilience and postoperative recovery.

The present study aimed to: (1) assess the degree to which unscheduled CD, compared to other delivery modes and accounting for prepartum and peripartum factors, is associated with clinically significant stress responses to childbirth, (2) determine whether these acute responses endure into the early postpartum period and (3) predict subsequent mental health symptoms.

## Materials and Methods

### Study Population

A total of 1,146 women who recently gave birth and received routine perinatal care at a tertiary, urban hospital, and completed an immediate postpartum assessment for acute stress were included in this study. Of these women, 340 (29.7%) underwent cesarean delivery and 806 (70.3%) delivered vaginally. Participants were assessed on average 31 hours post-delivery (range= 0.4 to 120 hours), primarily through in-person evaluations conducted during their maternity hospitalization. A subgroup (n=795, 69.4%) completed a second assessment approximately 1.92 months postpartum (range= 0.6 to 2.67). Participants were assessed during the third trimester for mental health and prior trauma; obstetrical data were extracted from electronic medical records.

The sample is derived from two prospective, longitudinal studies on mental health sequelae and childbirth experiences. Participants were third-trimester patients who planned to deliver at the study hospital. Recruitment for Study 1 occurred between October 2016 and October 2022 during routine perinatal visits. Recruitment for Study 2 occurred from March 2023 with data available through January 2025 and via hospital’s Patient Gateway portal. Both studies were approved by the hospital’s Human Research Committee. Participants provided implied consent by completing the study questionnaires via Redcap after receiving detailed information about study procedures.

## Measures

### Mental health

^111,2^Acute stress reactions to childbirth were assessed using the Peritraumatic Distress Inventory (PDI).^31^ This is a 13-item questionnaire that measures emotional and physiological responses experienced during and shortly after a traumatic event.^31^ Responses are rated on a 0 (Not at all) to 4 (Extremely true) scale with higher total scores indicating greater distress. The PDI demonstrates strong reliability and validity across trauma-exposed populations and postpartum samples;^21–23,36–39^ and clinical cutoff scores inform PTSD risk^27,29–31,40^ A cutoff score of 15 (excluding item 4) has been suggested to identify individuals at risk for childbirth-related PTSD^41^ and was used in the present study. Reliability of the PDI was high (α=0.83).

Depression symptoms were assessed using the Edinburgh Postnatal Depression Scale (EPDS), the recommended screening tool for peripartum depression in clinical settings.^40^ EPDS scores were primarily obtained from participants’ medical records. A score of 13 or higher was used as the clinical cutoff indicating probable depression.^42–45^

Posttraumatic stress disorder symptoms were measured with the well-validated PTSD Checklist for DSM-5 (PCL-5) a 20-item self-report questionnaire that assesses DSM-5 PTSD symptoms.^46–47^ Items are rated on a scale from 0 (Not at all) to 4 (Extremely); higher total scores reflect greater symptom severity. The PCL-5 demonstrates strong validity,^47–48^ including in perinatal populations.^49–50^ A cutoff of 32 is used for a provisional PTSD diagnosis.^47,49–51^ Reliability was high (for pregnancy α= 0.93; for postpartum assessment specified to recent childbirth: α=0.91).

Maternal-infant attachment problems were measured with the Maternal Attachment Inventory (MAI)^84^. This is a 26-item self-report questionnaire designed to assess maternal attachment with the infant and aligns with observational assessments. Items are rated on a 4- point scale, with higher scores indicating stronger perceived attachment impairment (α= 0.92).

Trauma history was assessed using the Life Events Checklist for DSM-5 (LEC-5),^53^ which captures exposure to various traumatic events (e.g., natural disasters, physical or sexual assault, sudden accidental death). The total number of events endorsed as “happened to me” was summed to generate a total trauma exposure score.

### Obstetric information

Delivery mode was categorized as vaginal, assisted vaginal, scheduled (elective) cesarean, and unscheduled cesarean (including emergent cases based on physician’s indication). Severity of obstetric complications was classified as: severe, life-threatening conditions per the American College of Obstetrics and Gynecology (ACOG)^54^ (e.g., uterine rupture, eclampsia, unplanned hysterectomy); moderate, significant morbidity or long-term risk (e.g., fetal labor intolerance, hemorrhage ≥ 1500cc, blood transfusion, 3rd/4th-degree lacerations, and infant health complications); mild, short-term complications (e.g., manual placenta removal without hemorrhage, failure to progress); or none. Additional medical records data included gestational week at delivery, primiparity, labor induction, and epidural administration.

### Analysis

Missing data for single items for mental health measures (i.e., PDI, PCL-5, EPDS, MAI for 3.04% of the mental health data) and other factors (i.e., demographics, trauma history, and obstetrical data for 3.85% of this data) (3.15% of the total data) were determined to be missing completely at random (MCAR) (Little’s MCAR Test, χ²(10901)=4548.0, p=1), and were handled with multiple imputation using the ‘mice’ package in R.^55^ Descriptive statistics were computed using proportions for categorical variables and means with standard deviations for continuous variables. Differences in rates of significant acute stress (PDI≥15) and background, obstetrics and mental health data were evaluated across delivery modes using chi-square tests of independence (or Fisher’s exact test) and analysis of variance performed for continuous variables. Comparisons for acute stress by delivery mode included adjusted relative risks and were computed using log-binomial regression, controlling for demographic, mental health and obstetric factors, if associated with PDI score (p≤0.10). Repeated measures linear mixed model with Kenward-Roger adjustment was used to assess stability of stress levels between time points and across delivery modes adjusting for identified contributors with planned post-hoc analyses using estimated marginal means (EMMs) and Sidak adjustment to identify between and within-group changes. We also examined stress rates stratified by severity of obstetric complications (and labor stage for unscheduled CD) and used regression analysis to determine the prediction of acute stress of later mental health outcomes controlling for obstetric factors. Statistical analyses were conducted in R (version 4.3.0).^56^

## Results

The mean maternal age was 33.7 (SD=4.07) years; 38.4% of participants were primiparous. Most women (95%) delivered at full-term and 70.3% had vaginal deliveries, while 29.7% underwent CD. Obstetric complications were identified in 36.9% of cases with 0.2% life-threatening complications. Table 1 presents sample demographics, prepartum mental health status, and obstetric information, stratified by delivery mode.

**Table 1.**
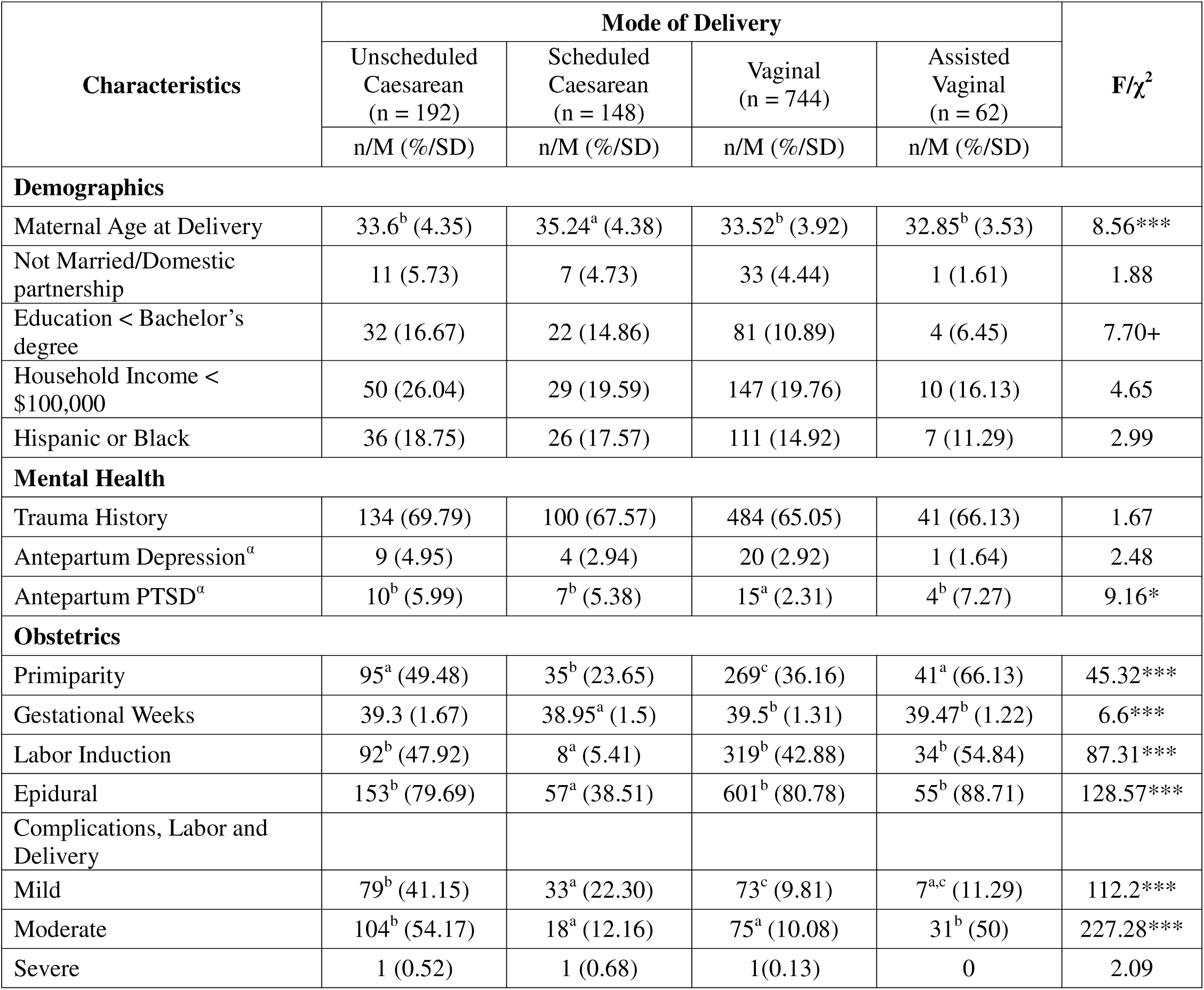
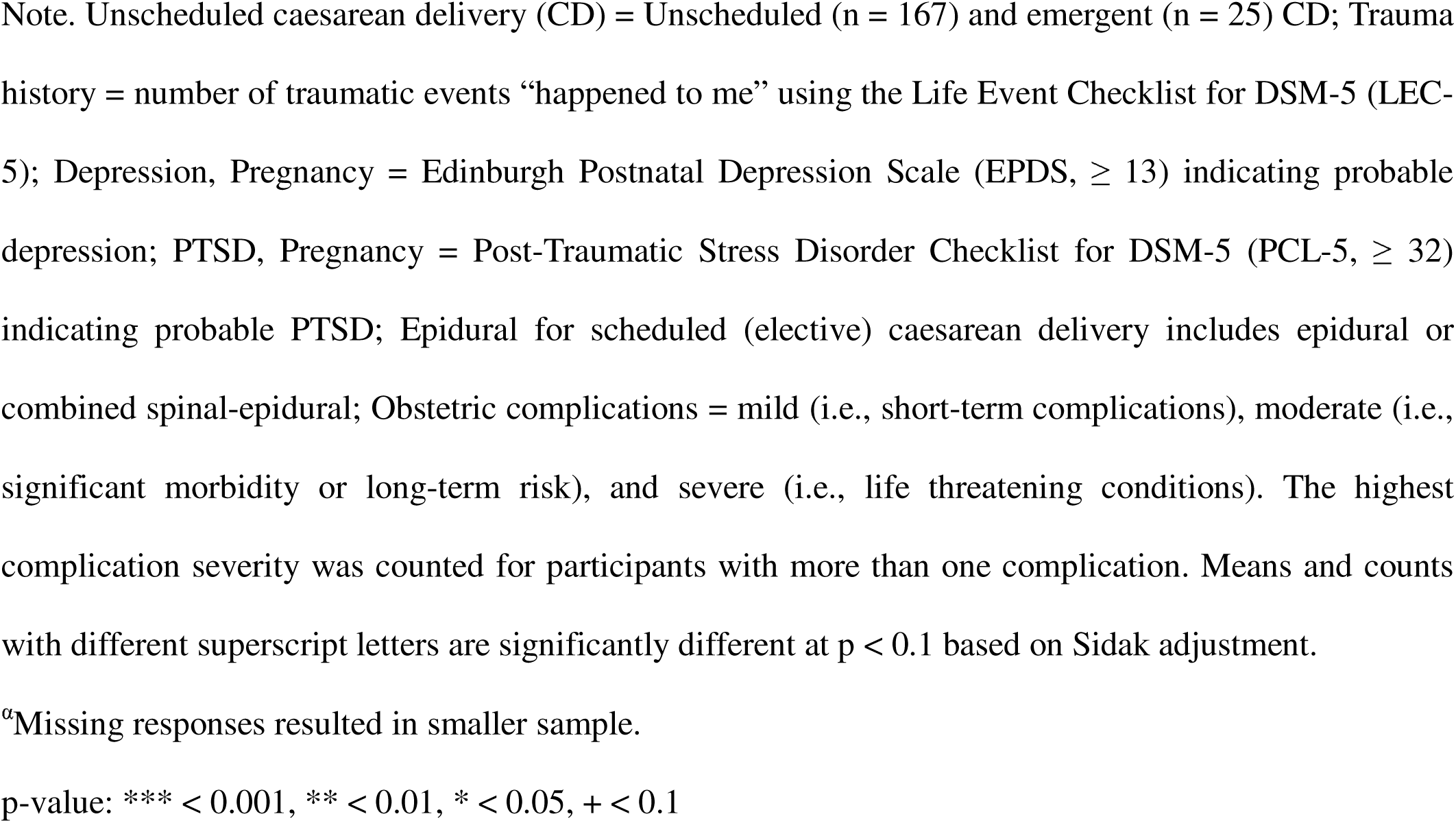
Demographics, mental health, and obstetric factors by mode of delivery.

At the immediate postpartum assessment (∼31 hours postpartum), 10.5% (n=120) of the sample reported clinical stress (PDI≥15) occurring during childbirth or immediately after. At the later timepoint (∼1.9 month postpartum), 9.6% (n=76) continued to meet this threshold. Stress responses associated with the obstetric complication severity (ρ=0.28, p<0.001) (Supplementary Table 1).

### Acute Stress Reactions to Childbirth and Delivery Mode

Among participants who underwent unscheduled CD, 26.6% endorsed clinically significant acute stress reactions. Rates varied by stage of labor during which the unscheduled CD was performed: 14.3% when non-laboring (5/35), 25.3% in the first stage (23/91), and 34.9% during the second stage (23/66) (Table 2). Rates were higher for unscheduled CD during laboring in comparison to non-laboring (χ² (1)=3.31, p=0.07) and stress levels were correlated with obstetric complications severity, ρ= 0.20, p= 0.005 (20.3% and 33.7% in patients with minor and moderate complications). In contrast, acute stress rates were 6.3% among participants with vaginal delivery and 22.6% in assisted vaginal delivery (28.6% and 29.0% in minor/ moderate obstetric complications) (Table 3).

**Table 2.** Acute stress response by labor stage and obstetric morbidity in unscheduled caesarean delivery.

| Labor Stage | Complication Severity | Acute Stress<br>(PDI ≥ 15) |
| --- | --- | --- |
|  |  | n (%) |
| Non-laboring | None | 0 |
|  | Mild | 3 (15.79) |
|  | Moderate | 2 (14.29) |
|  | Severe | - |
| First Stage | None | 0 |
|  | Mild | 4 (14.29) |
|  | Moderate | 19 (33.33) |
|  | Severe | - |
| Second Stage | None | - |
|  | Mild | 9 (28.13) |
|  | Moderate | 14 (42.42) |
|  | Severe | 0 |

Note. Acute stress reactions to recent childbirth measured in the immediate postpartum (~31hrs) using the Peritraumatic Distress Inventory (PDI), clinical cutoff ( $\geq 15$ ); Labor stages during performance of unscheduled Cesarean delivery = non-laboring (n = 35): prior to initiation of labor; first stage (n = 91): from onset of labor to full cervical dilation; and second stage (n = 66): full dilation to fetal delivery; Obstetric complications severity = mild: short-term complications; moderate: significant morbidity or long-term risk, including infant health
complications; and severe: life-threatening conditions. For multiple complications, the highest severity of complication was counted. n = 192.

**Table 3.**
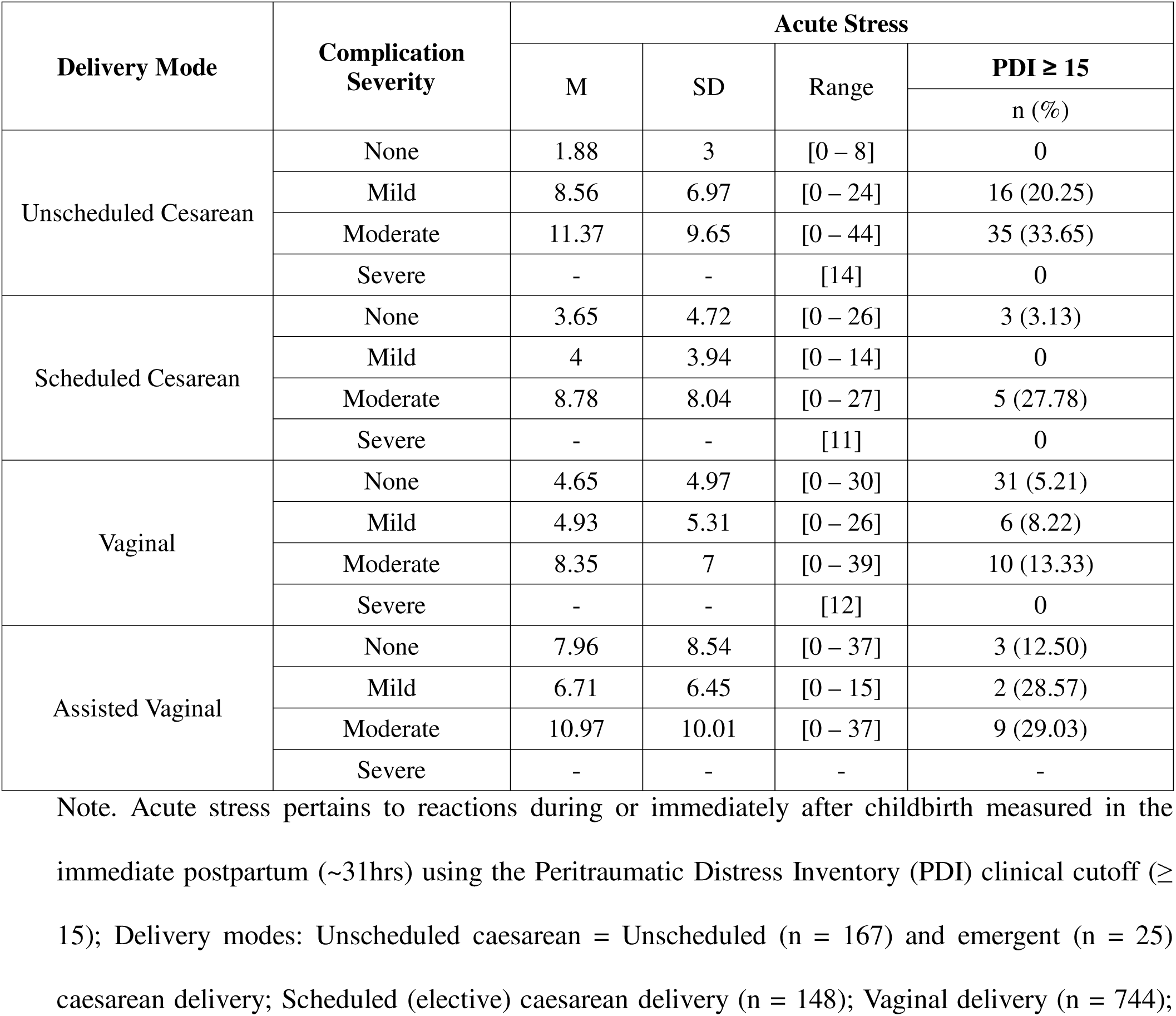
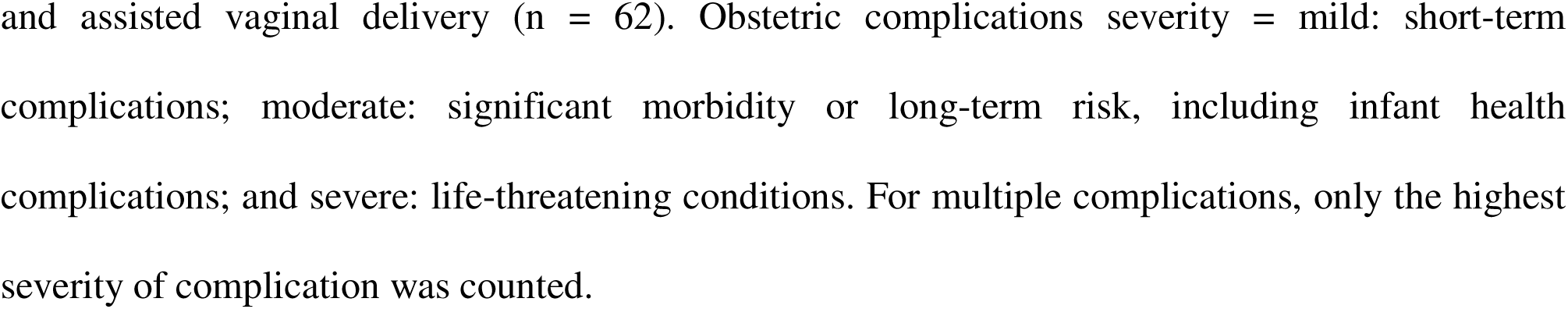
Acute stress response by delivery mode and obstetric morbidity.

| Delivery Mode | Complication Severity | Acute Stress |  |  |  |
| --- | --- | --- | --- | --- | --- |
|  |  | M | SD | Range | PDI ≥ 15 |
|  |  |  |  |  | n (%) |
| Unscheduled Cesarean | None | 1.88 | 3 | [0 – 8] | 0 |
|  | Mild | 8.56 | 6.97 | [0 – 24] | 16 (20.25) |
|  | Moderate | 11.37 | 9.65 | [0 – 44] | 35 (33.65) |
|  | Severe | - | - | [14] | 0 |
| Scheduled Cesarean | None | 3.65 | 4.72 | [0 – 26] | 3 (3.13) |
|  | Mild | 4 | 3.94 | [0 – 14] | 0 |
|  | Moderate | 8.78 | 8.04 | [0 – 27] | 5 (27.78) |
|  | Severe | - | - | [11] | 0 |
| Vaginal | None | 4.65 | 4.97 | [0 – 30] | 31 (5.21) |
|  | Mild | 4.93 | 5.31 | [0 – 26] | 6 (8.22) |
|  | Moderate | 8.35 | 7 | [0 – 39] | 10 (13.33) |
|  | Severe | - | - | [12] | 0 |
| Assisted Vaginal | None | 7.96 | 8.54 | [0 – 37] | 3 (12.50) |
|  | Mild | 6.71 | 6.45 | [0 – 15] | 2 (28.57) |
|  | Moderate | 10.97 | 10.01 | [0 – 37] | 9 (29.03) |
|  | Severe | - | - | - | - |

Note. Acute stress pertains to reactions during or immediately after childbirth measured in the immediate postpartum (~31hrs) using the Peritraumatic Distress Inventory (PDI) clinical cutoff (≥ 15); Delivery modes: Unscheduled caesarean = Unscheduled (n = 167) and emergent (n = 25) caesarean delivery; Scheduled (elective) caesarean delivery (n = 148); Vaginal delivery (n = 744);
and assisted vaginal delivery (n = 62). Obstetric complications severity = mild: short-term complications; moderate: significant morbidity or long-term risk, including infant health complications; and severe: life-threatening conditions. For multiple complications, only the highest severity of complication was counted.

Table 4 presents risk for acute stress by delivery mode. The rate of clinically significant acute stress was higher in response to unscheduled CD, compared to other modes (excluding assisted vaginal delivery) (χ² (3)=80.47, p< 0.001). At the immediate postpartum assessment, the relative risk (RR) of acute stress in unscheduled CD, compared to vaginal delivery, was 4.20 (CI: [2.92–6.05]). Accounting for contributors associated with acute stress levels (p<=0.10) (i.e., maternal age, education, income, primiparity, antenatal depression/PTSD, induction and obstetric complications), adjusted relative risk (aRR) was 2.80 (CI: [1.72–4.55]). The unscheduled CD group also had a higher risk for acute stress, compared to scheduled cesarean (RR=4.91, CI: [2.41–10.03], aRR=3.52, CI: [1.47–8.44]).

**Table 4.** Acute stress responses to childbirth by mode of delivery.

| Mode of Delivery | Acute Stress |  |  |  |  |  |
| --- | --- | --- | --- | --- | --- | --- |
| | PDI ≥ 15 | $\chi^2$ | Relative Risk | M | SD | Range |
|  | n (%) |  |  |  |  |  |
| Unscheduled Caesarean | 51 (26.56) | 80.47*** | - | 9.83 | 8.66 | [0 – 44] |
| Scheduled Caesarean | 8 (5.41) |  | 4.91*** [2.41 – 10.03]<br>3.52** [1.47 – 8.44]<sup>a</sup> | 4.40 | 5.32 | [0 – 27] |
| Vaginal | 47 (6.32) |  | 4.20*** [2.92 – 6.05]<br>2.80*** [1.72 – 4.55]<sup>a</sup> | 5.06 | 5.35 | [0 – 39] |
| Assisted Vaginal | 14 (22.58) |  | 1.25 [0.65 – 2.41]<br>1.15 [0.70 – 1.91]<sup>a</sup> | 9.32 | 9.15 | [0 – 37] |

Note. Patient assessment completed ~31 hours after childbirth (n = 1,146). Stress response measured using peritraumatic distress inventory (PDI) clinical cutoff ≥ 15. Unscheduled caesarean delivery (CD) = Unscheduled (n = 167) and emergent (n = 25) CD; Scheduled (elective) caesarean delivery (n = 148); vaginal delivery (n = 744); and assisted vaginal delivery (n = 62).
<sup>a</sup>Adjusted relative risk in reference to unscheduled CD accounting for maternal age, education, income, trauma history, antepartum depression and PTSD, primiparity, labor induction and obstetric complications.
p-value: \*\*\* < 0.001, \*\* < 0.01, \* < 0.05, + < 0.1

Commonly endorsed acute stress items for women who had unscheduled CD included having physical stress reactions (64.6%), feeling helpless (37%) and as though they might pass out (37%), and thoughts of dying (14.6%) (Figure 1).

**Figure 1.**
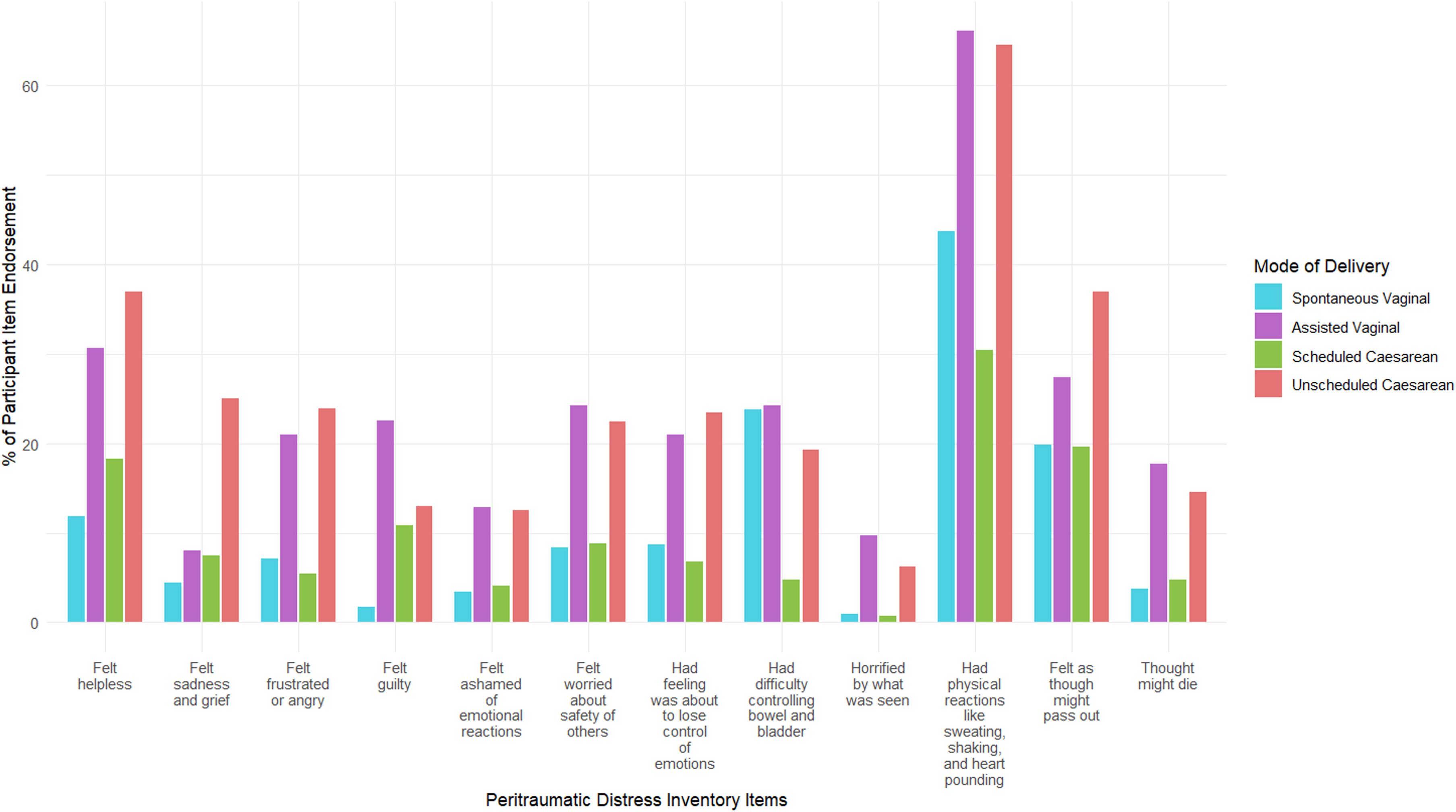
Characteristics of acute stress responses to childbirth by mode of delivery. The histogram bars represent percentage of participants by delivery mode with endorsement of a score of 2 or higher (i.e., at least to a moderate degree) on each of the 12 items of the Peritraumatic Traumatic Distress Inventory (PDI). Assessment obtained ∼31 hours after childbirth. n = 1,146.

### Stability of Acute Stress and Delivery Mode

Figure 2 presents acute stress levels in response to childbirth by delivery mode assessed across time points (Model fit, Marginal R²=0.22; Conditional R²=0.73). Significant fixed effects were observed for delivery mode (F(3, 662)=13.47, p<0.001) and for the interaction (delivery by time) (F(3, 671)=3.47, p=0.02. The time effect was not significant (F(1, 671)=0.45, p=0.50).

**Figure 2.**
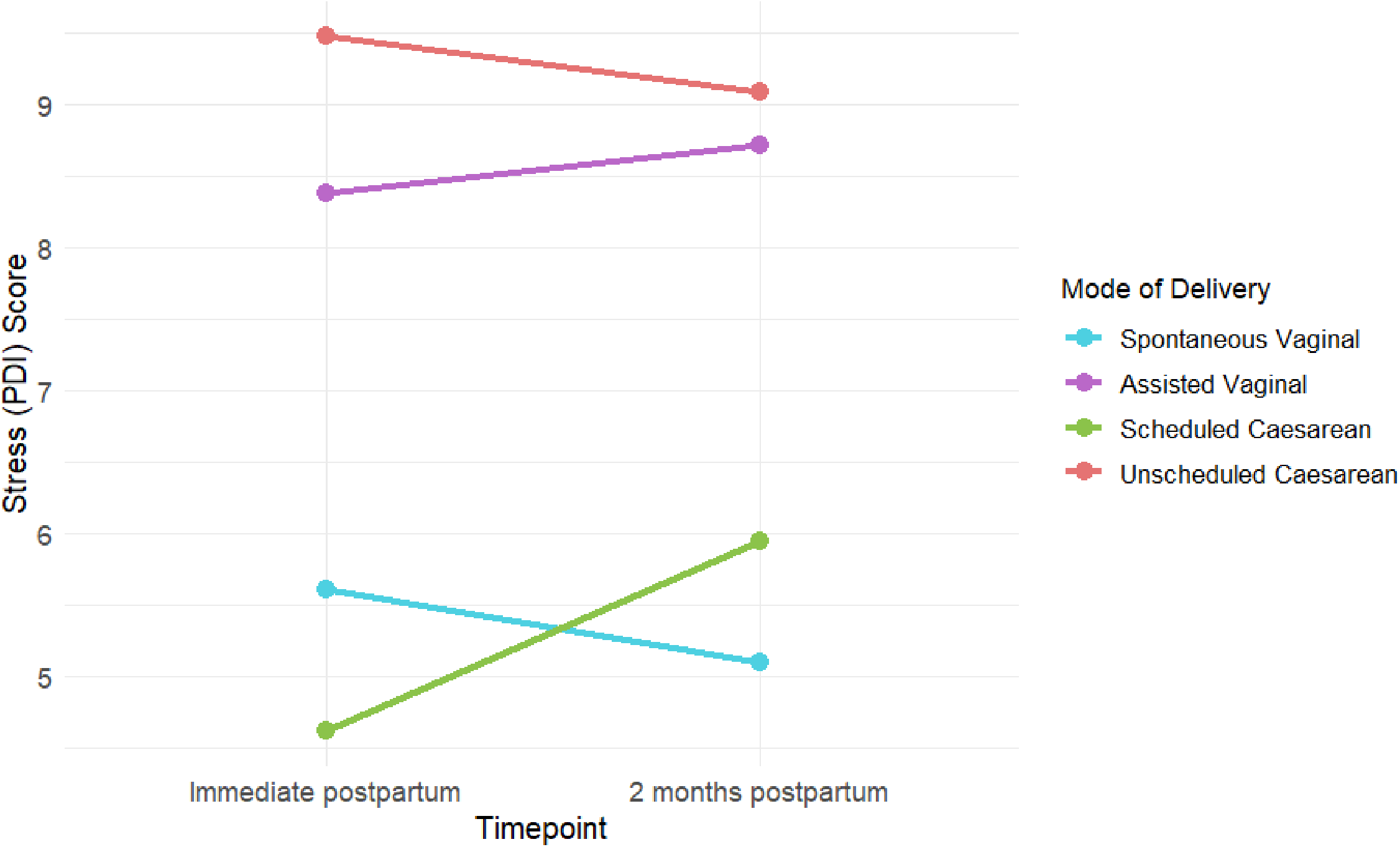
Level of acute stress to childbirth over time by delivery mode. The figure depicts stress levels in response to recent childbirth assessed by the Peritraumatic Distress Inventory (PDI) (total scores) for each delivery mode in the immediate postpartum assessment (∼29.4 hours post-delivery) time point and the second (∼1.9 months post-delivery) assessment. n = 795.

Planned contrasts, with unscheduled CD as reference group, showed that the unscheduled CD group had higher stress (PDI score) across time points in comparison to vaginal delivery (estimate=3.93, p<0.001) and scheduled cesarean (estimate=4.00, p<0.001) but not assisted vaginal delivery (estimate= 0.73, p= 0.87). While stress levels were stable for the unscheduled CD (estimate=0.39, p=0.44) and assisted vaginal delivery (estimate=0.34, p=0.68), the levels decreased for vaginal delivery (estimate=0.52, p=0.03) and increased for scheduled cesarean (estimate= 1.33, p= 0.01) accounting for contributing factors.

### Acute Stress as a Risk Factor for Later Mental Health Problems

Regression analyses revealed that acute stress reported immediately postpartum significantly predicted mental health and maternal-infant bonding outcomes at two months. Accounting for obstetric factors (i.e., delivery mode, complications severity, primiparity, epidural use and induction), acute stress levels predicted PTSD symptoms (β=0.48, p<0.001, R²-change, ΔR²=0.19) depressive symptoms (β= 0.30, p<0.00, ΔR²=0.09) and bonding difficulties (β=0.32, p<0.001, ΔR²=0.08).

## Discussion

### a. Principal Findings

The reported study documents high rates of psychological stress among patients who underwent unscheduled CD, most of whom experienced non-emergent procedures. 26.6% of unscheduled CD patients reported clinically significant acute stress reactions occurring during childbirth or immediately after with higher rates related to stages of labor (29.3% in CD performed in labor) and obstetric morbidity. The risk of endorsing acute stress was approximately three times higher for patients undergoing unscheduled CD, compared to those with vaginal delivery, after accounting for factors including demographic background, primiparity, obstetric complications, labor induction, antepartum mental health status, and trauma history. Reported heightened traumatic stress among women with unscheduled CD persisted through the early postpartum months, while those who had vaginal delivery exhibited low stress levels that further declined over time. Acute stress responses were strongly associated with subsequent PTSD and depressive symptoms and maternal-infant bonding problems. These findings demonstrate that unscheduled CD can signal increased vulnerability to psychological stress during childbirth and heightened stress may have enduring effects on maternal mental health. Importantly, rates of acute stress were similar between women undergoing unscheduled CD and those with assisted vaginal delivery, suggesting that underlying obstetric acuity, the need for an unanticipated intervention, or morbidity might be associated with stress responses, rather than the procedure itself.

### b. Results in the Context of What is Known

There is growing recognition of childbirth as an experience that can be psychologically stressful for some individuals.^57–60^ Prior studies have shown that unscheduled CD is associated with an increased risk of developing PTSD symptoms.^36,61–64^ Yet, there remains limited understanding of the immediate psychological impact of unscheduled CD. This study documents significant stress reactions during childbirth in women who undergo unscheduled CD. These immediate reactions are clinically meaningful, as they may serve as early indicators of subsequent maternal posttraumatic and depressive symptoms and maternal-infant attachment problems. The pattern of clinically significant stress we observed closely mirrors immediate psychological responses reported in individuals exposed to other traumas.^29–31^

### c. Clinical Implications

While the first 24 hours after childbirth is well recognized as a critical window for monitoring physical recovery,^54,65–68^ especially after cesarean delivery,^69^ the psychological contribution of the mode of delivery has received limited attention in clinical practice and maternity care planning. This study demonstrates that unscheduled CD associates with heightened psychological stress reactions during this important period of maternal behavior initiation. Because peritraumatic reactions can strongly influence posttraumatic mental health trajectories,^25,32,33,41^ timely screening for acute stress responses, especially immediately following unscheduled CD or other medically complicated deliveries, would improve the accuracy of mental health determinations, enhance opportunities for early and effective treatment,^70^ and better prepare patients for future pregnancies.^24,41,70^

Currently, U.S. hospital screening protocols focus on maternal depression (e.g., using the Edinburgh Postnatal Depression Scale, EPDS) and do not assess traumatic stress. Our findings support the American College of Obstetricians and Gynecologists (ACOG) recommendation for more comprehensive mental health screening.^71^ Importantly, our findings support the potential utility of the PDI as a feasible and scalable screening tool that can be administered in the immediate postpartum for identifying individuals at risk for adverse mental health outcomes following complicated childbirth.^41^ Anticipating risk for traumatization during pregnancy may inform shared decision-making around birth plans and delivery mode and optimize maternal and neonatal outcomes.^72^

Unscheduled CD may disrupt oxytocin and cortisol surges that support labor.^73–74^ Dysregulated stress responses in combination with the threat of undergoing an unplanned major obstetric procedure and its acuity may increase susceptibility for traumatization. Acute stress reactions were higher among women who underwent unscheduled CD after labor had already started, compared to those who had unscheduled CD before labor began or scheduled cesarean delivery. We also found that acute stress levels were positively associated with degree of obstetric complications, suggesting a dose-response relationship between obstetric morbidity and acute stress. Thus, the disruption of naturally occurring maternal-stress regulation during labor and the events leading to the surgery may contribute to an increased acute stress response.

### d. Research Implications

An important direction for future research is the development of comprehensive risk assessment models that integrate obstetric factors commonly associated with unscheduled CD such as labor induction, pain intensity, epidural use, time of birth, and maternal or neonatal complications alongside pre-existing psychological vulnerabilities. Given that the subjective childbirth experience plays a key role in posttraumatic maternal outcomes,^75–76^ future studies are needed to investigate the influence of the presence of support person, staff support and patient-clinician interactions in shaping trauma responses. Understanding how these interpersonal and contextual factors amplify, or buffer distress will be essential for developing evidence-based recommendations for trauma-informed care. Gaining insight into the potential for psychological growth following a traumatic childbirth experience^79–80^ as well as the processes facilitating meaning-making may provide pathways for fostering maternal resilience.^77–78^

### e. Strengths and Limitations

A key strength of this study is the ability to assess patients’ stress reactions to childbirth during the immediate postpartum period. We used a well-established structured instrument to measure traumatic stress, previously validated in trauma-exposed populations, to capture clinically significant acute reactions to childbirth. Rather than relying on a community sample, the study was performed in a clinical setting to estimate rates of stress reaction. We conducted repeated assessments to evaluate the stability of recalled stress reactions and their association with postpartum mental health. The risk of acute stress associated with unscheduled CD was compared to other delivery modes. We incorporated electronic medical record data to assess obstetric complications, labor induction and important antepartum mental health status and prior trauma, which allowed more nuanced and adjusted analysis of unscheduled CD’s link with traumatic stress.

Several limitations should be noted. The study design does not permit the separation of the contribution of stressful or acute events leading to unscheduled CD from the contribution of the procedure itself to acute stress responses. It is thus not possible to know whether it is the unscheduled CD, or the reasons for the unscheduled CD, that are associated with increased acute stress responses compared to vaginal delivery. Women experiencing severe stress were potentially less likely to be approached, thereby, are less represented in the sample. We did not include biological measures of acute stress (e.g., heart rate variability), which could have provided mechanistic insights. Finally, although the obstetric characteristics of participants who completed both assessments were generally representative of the original cohort, a signification proportion was lost to follow-up. Cultural or sociodemographic differences may limit the generalizability of our findings to other populations or healthcare systems.

## Conclusions

Unscheduled CD can be lifesaving for both mother and fetus. Our findings provide evidence for an increased risk of psychological traumatization for patients who undergo unscheduled CD, with consequences for maternal mental health. Given that trauma exposure and stress can lead to profound neurobiological changes,^79–81^ and heighten vulnerability to re-traumatization with subsequent traumatic events,^33,82,83^ there is a need to prioritize birth trauma prevention in perinatal care. Patient and provider education is an essential first step that should facilitate implementation of trauma-informed screening, early intervention strategies, and systemic policy reforms.

## Declaration of generative AI and AI-assisted technologies in the writing process

During the preparation of this work the authors used ChatGPT-4o in order to improve language and readability and reduce word count. After using this tool/service, the authors reviewed and edited the content as needed and take full responsibility for the content of the publication.

## Data Availability

All data produced in the present study are available upon reasonable request to the authors

