## Supplementary Table 1 for "The psychological impact of childbirth: Unscheduled cesarean delivery increases risk for acute stress response"

Table 1. **Acute stress during childbirth by obstetric complications**

| **Complication Severity** |  | **n (%)** | **PDI score**  **Mean/Range** | **PDI≥15**  **N (%)** |
| --- | --- | --- | --- | --- |
| Mild | Hypertension^a^ without severe features | 17 (1.48) | 7.66 (0-22) | 5 (29.4) |
|  | Complications of anaesthesia | 1 (0.09) | 26 | 1 (100) |
|  | Chorioamnionitis^b^ | 32 (2.79) | 5.06 (0-20) | 4 (12.5) |
|  | PPH^c^ | 45 (3.92) | 6.13 (0-21) | 5 (11) |
|  | Removal of retained placenta without PPH | 23 (2) | 5.69 (0-18) | 2 (8.7) |
|  | Failed induction of labor | 1 (0.09) | 18 | 1 (100) |
|  | Failed TOLAC^d^ | 1 (0.09) | 16 | 1 (100) |
|  | Failure to progress in 1^st^/2^nd^ stage | 64 (5.58) | 9.07 (0-27) | 13 (20.3) |
|  | PROM^e^ | 11 (0.96) | 2.9 (0-9) | 0 |
|  | Malpresentation | 28 (2.44) | 5.82 (0-21) | 3 (19.7) |
|  | Placental abruption without PPH | 6 (0.52) | 3.67 (6-0) | 0 |
|  | Precipitous delivery^f^ | 1 (0.09) | 6 | 0 |
| Moderate | PPH^c^ ≥ 1500 mL | 20 (1.75) | 12.35 (0-44) | 6 (30) |
|  | Need for blood transfusions | 12 (1.04) | 12.33 (0-37) | 3 (25) |
|  | 3^rd^/4^th^ perineal lacerations degree | 27 (2.35) | 8.29 (0-28) | 3 (11) |
|  | Removal of retained placenta with PPH | 5 (0.44) | 13 (0-30) | 2 (40) |
|  | Hypertension^a^ with severe features | 8 (0.69) | 2 (0-9) | 0 |
|  | Shoulder dystocia | 12 (1.04) | 11 (0-39) | 2 (16.7) |
|  | Hysterotomy extension to the vagina | 3 (0.26) | 8 (6-10) | 0 |
|  | Fetal intolerance of labor | 109 (9.51) | 11.48 (0-44) | 36 (33) |
|  | Cord prolapse | 1 (0.09) | 0 | 0 |
|  | Failed vacuum/forceps | 2 (0.17) | 5.5 (3-8) | 0 |
|  | Neonatal complications^g^ | 92 (8.55) | 10.23 (0-37) | 25 (27.1) |
| Severe | ACOG Criteria | 3 (0.26) | 12.33 (11-14) | 0 |

Note. Mild, moderate and severe complications defined as short-term complications, significant morbidity or long-term risk, or life-threatening conditions per American College of Obstetrics and Gynecology (ACOG), respectively. Mild complications count (n) includes only cases without moderate complications. a- Pre-eclampsia or gestational hypertension; b- Chorioamnionitis, intraamniotic infection, or maternal T>100.4; c- PPH (Post-partum hemorrhage) based on EBL (estimated blood loss), mild: 1000 ml £EBL <1500 ml without blood transfusion or hemorrhage noted in medical records in cases of EBL <1000 ml, moderate, EBL ³1500 ml or placing of a uterine balloon or uterine compression suture and £1 unit of blood products transfused; d- TOLAC (trail of labor after Caesarean); e- PROM (premature rupture of membranes); f- Precipitous delivery without epidural anaesthesia; g- Neonetal complications- Prematurity, 5-minute APGAR score <7, Neonatal Intensive Care Unit (NICU) admission.
